# Infectivity and immune escape of the new SARS-CoV-2 variant of interest Lambda

**DOI:** 10.1101/2021.06.28.21259673

**Authors:** Mónica L. Acevedo, Luis Alonso-Palomares, Andrés Bustamante, Aldo Gaggero, Fabio Paredes, Claudia P. Cortés, Fernando Valiente-Echeverría, Ricardo Soto-Rifo

## Abstract

**Background:** The newly described SARS-CoV-2 lineage C.37 was recently classified as a variant of interest by the WHO (Lambda variant) based on its high circulation rates in South American countries and the presence of critical mutations in the spike protein. The impact of such mutations in infectivity and immune escape from neutralizing antibodies are entirely unknown.

**Methods:** We performed a pseudotyped virus neutralization assay and determined the impact of the Lambda variant on infectivity and immune escape using plasma samples from healthcare workers (HCW) from two centers in Santiago, Chile who received the two-doses scheme of the inactivated virus vaccine CoronaVac.

**Results:** We observed an increased infectivity mediated by the Lambda spike protein that was even higher than that of the D614G (lineage B) or the Alpha and Gamma variants. Compared to the Wild type (lineage A), neutralization was decreased by 3.05-fold for the Lambda variant while it was 2.33-fold for the Gamma variant and 2.03-fold for the Alpha variant.

**Conclusions:** Our results indicate that mutations present in the spike protein of the Lambda variant of interest confer increased infectivity and immune escape from neutralizing antibodies elicited by CoronaVac. These data reinforce the idea that massive vaccination campaigns in countries with high SARS-CoV-2 circulation must be accompanied by strict genomic surveillance allowing the identification of new isolates carrying spike mutations and immunology studies aimed to determine the impact of these mutations in immune escape and vaccines breakthrough.

---

The emergence of SARS-CoV-2 variants of concern and variants of interest has been a hallmark of the COVID-19 pandemic during 2021^1–3^. The newly assigned SARS-CoV-2 lineage C.37 was recently classified as a variant of interest by the WHO on June 14^th^ and denominated as the Lambda variant ^4^. The presence of this new variant has been reported in more than 20 countries as per June 2021 with most of the available sequences coming from South American countries, particularly from Chile, Peru, Ecuador and Argentina^5^. This new variant of interest is characterized by the presence of a convergent deletion in the ORF1a gene (Δ3675-3677) already described in the Beta and Gamma variants of concern and mutations Δ246-252, G75V, T76I, L452Q, F490S, T859N in the spike protein^6^. The impact of these spike mutations on infectivity and escape to neutralizing antibodies are completely unknown.

Chile is currently undergoing a massive vaccination program. According to public data from the Chilean Ministry of Health as per June 27^th^ 2021, 65.6% of the target population (18 years old and older) have received a complete vaccination scheme^7^. The vast majority (78.2%) of the fully vaccinated population have received the two doses scheme of the inactivated virus vaccine CoronaVac, which has been previously reported to elicit neutralizing antibodies but at lower titers when compared to plasma or sera from convalescent individuals^8–11^.

Here, we used our previously described pseudotyped virus neutralization assay^12^ to determine the impact of the Lambda variant on the neutralizing antibodies responses elicited by the inactivated virus vaccine CoronaVac. Our data show that mutations present in the spike protein of the Lambda variant confer increased infectivity and escape to neutralizing antibodies elicited by the inactivated virus vaccine CoronaVac.

## Methods

### Study cohort

Health care workers from two sites in Santiago, Chile were invited to participate. Volunteers received the two-doses scheme of CoronaVac, each dose being administered 28 days apart according to the Chilean vaccination program. Plasma samples were collected between May and June 2021. All participants signed an informed consent before any study procedure was undertaken.

### Public data analysis

Data on SARS-CoV-2 lineages and the date the sample was taken from the sequences available from Chile were obtained from the Consorcio Genomas CoV2 site available at https://auspice.cov2.cl/ncov/chile-global. Vaccination data were obtained from public data from Ministry of Science, Technology, Knowledge and Innovation available at https://github.com/MinCiencia/Datos-COVID19 (Product 83).

### Infectivity assay

Pseudotyped viruses carrying different SARS-CoV-2 spike proteins were prepared as we previously described^12^. Briefly, HIV-1-based SARS-CoV-2 pseudotypes were produced in HEK293T cells by transfecting the pNL4.3-ΔEnv-Luc together with the corresponding pCDNA-SARS-CoV-2 Spike coding vector in a 1:1 molar ratio. Plasmids coding a codon-optimized spike lacking the last 19 amino acids of the C-terminal end (SΔ19) known to avoid retention at the endoplasmic reticulum^12^ were obtained by gene synthesis or customized site-directed mutagenesis (GeneScript) and contained the following mutations: lineage A (reference sequence), lineage B (D614G), lineage B.1.1.7 (Δ69-70, Δ144, N501Y, A570D, D614G, P681H, T716I, S982A, D1118H), lineage P.1 (L18F, T20N, P26S, D138Y, R190S, K417T, E484K, N501Y, D614G, H655Y, T1027I) and lineage C.37 (G75V, T76I, Δ246-252, L452Q, F490S, D614G, T859N). Each pseudotype preparation was cleared by centrifugation at 3,000 rpm at room temperature, quantified using the HIV-1 Gag p24 Quantikine ELISA Kit (R&D Systems), aliquoted in 50% fetal bovine serum (Sigma-Aldrich) and stored at -80 °C until use. Different amounts of pseudotyped viruses (as determined by the levels of the HIV-1 p24 protein) were used to infect HEK-ACE2 cells and 48 hours later, firefly luciferase activity was measured using the Luciferase Assay Reagent (Promega) in a Glomax 96 Microplate luminometer (Promega).

### Neutralization assay

Pseudotyped virus neutralization assays were performed essentially as we previously described^12^. Briefly, serial dilutions of the plasma samples (1:4 to 1:8748) were prepared in DMEM with 10% fetal bovine serum and incubated with 5 ng of p24 of each pseudotyped virus during 1h at 37 °C and then, 1×10^4^ of HEK-ACE2 cells were added to each well. HEK293T cells (not expressing ACE2) incubated with the pseudotyped virus (lineage A) were used as a negative control. Cells were lysed 48 hours later, and firefly luciferase activity was measured using the Luciferase Assay Reagent (Promega) in a Glomax 96 Microplate luminometer (Promega). The percentage of neutralization for each dilution was calculated and the ID50 of each sample was calculated using GraphPad Prism version 9.0.1.

### Statistical analyses

Statistical analyses were performed using GraphPad Prism software version 9.1.2. Multiple group comparisons for neutralizing antibody titers (NAbTs) against a panel of SARS-CoV-2 pseudotyped viruses as well as comparisons of NAbs responses by sex and smoke status were carried out using a paired Wilcoxon signed-ranked test. Factor change was calculated as the difference of geometric mean titer in the ID_50_ as compared with that of the Wild type pseudotyped virus. Correlation analysis between NAbTs and age or BMI was carried out using Spearman’s test. One-way ANOVA and Tukey’s multiple comparison test were performed for statistical analysis of infectivity. A p value ≤0.05 was considered as statistically significant.

### Ethical approval

The study protocol was approved by the Ethics Committee of the Faculty of Medicine at Universidad de Chile (Projects N° 0361-2021 and N° 096-2020) and Clínica Santa Maria (Project N°132604-21). All donors signed the informed consent, and their samples were anonymized.

## RESULTS

### Impact of the spike mutations in the Lambda variant on infectivity and neutralizing antibodies responses

An analysis of 3695 sequences from Chile deposited at GISAID as per June 24^th^ 2021 shows a clear dominance of the SARS-CoV-2 variants Gamma and Lambda during the last trimester accounting, together, for the 79% of all sequences (Appendix Figure 1). Interestingly, this period has been characterized by a massive vaccination campaign in which 65.6% of the target population (18 years old and older individuals) have received a complete vaccination scheme as per June 27^th^ 2021^7^. Given that 78.2% of the people inoculated with a complete scheme received the inactivated virus vaccine CoronaVac from Sinovac Biotech (Appendix Figure 1), we sought to investigate the impact of the spike mutations present in the Lambda variant on the neutralizing capacity of antibodies elicited by this vaccine. For this, we generated HIV-1-based SARS-CoV-2 pseudotyped viruses carrying the spike protein from the Wuhan-1 reference lineage (Wild type; lineage A), the D614G mutation (lineage B), and the Alpha (lineage B.1.1.7), Gamma (lineage P.1) and Lambda (lineage C.37) variants. During virus preparation, we consistently observed that cells infected with the pseudotyped virus carrying the Lambda spike produced significant higher bioluminescence values when compared to the D614G mutant or the Alpha and Gamma variants indicating an increased infectivity driven by the Lambda spike protein (Figure 1 and Appendix, Table 1).

**Figure 1.**
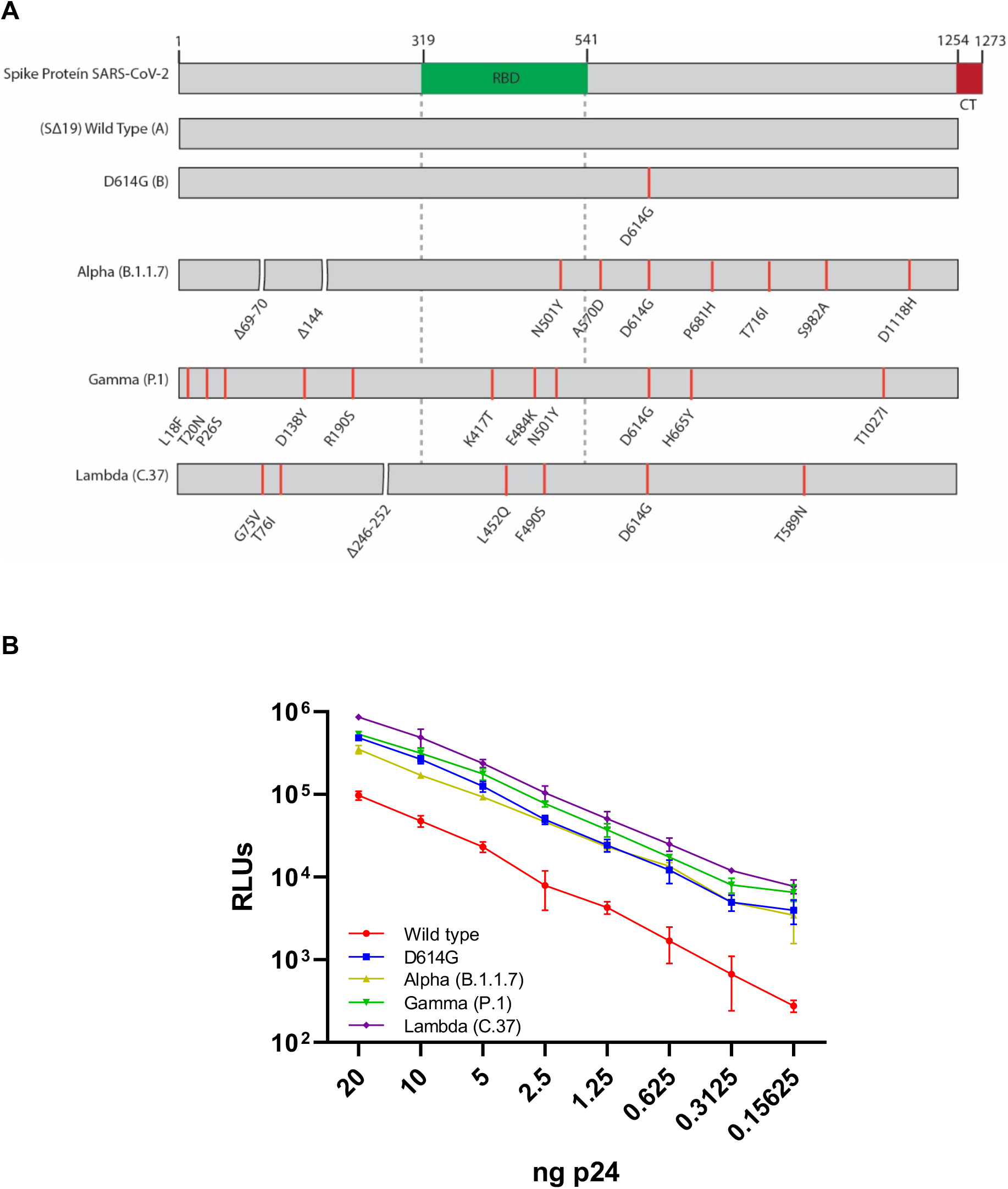
Infectivity mediated by different spike proteins. (A) Schematic representation of the SARS-CoV-2 spike protein and the variants used in this study. Lineages are indicated in parenthesis. RBD, receptor-binding domain, CM; cytoplasmic tail. (B) Titration of the pseudotypes of each lineage using equivalent amounts of HIV-1 p24. The firefly luciferase activity was measured as relative luminescence units (RLU) at 48 hours post-infection. The average and SD were calculated from a representative triplicate experiment. See Appendix Table 1 for statistical analyses.

Next, we used the pseudotyped viruses mentioned above to performed neutralization assays using 79 plasma samples from healthy healthcare workers from Universidad de Chile and Clínica Santa María at Santiago, Chile (Appendix, Table 2). We excluded 4 samples as we were not able to calculate an ID50 titer. From the analyzed samples, 73% corresponded to women, median age 34 years (IQR 29 – 43) and a body max index (BMI) of 25 (IQR 22.7 – 27). A 20.5% of the participants declared be an active smoker while the immunization period lasted. Samples were obtained in a median of 95 days (IQR 76 – 96) after the second dose of CoronaVac vaccine (Appendix, Table 2).

We observed that neutralization of the pseudotyped virus carrying the Wild type spike protein resulted in a 50% inhibitory dilution (ID_50_) mean titer of 191.46 (154.9 – 227.95, 95% CI,), while it was 153.92 (115.68 – 192.16, 95% CI), 124.73 (86.2 – 163.2, 95% CI), 104.57 (75.02 – 134.11, 95% CI) and 78.75 (49.8 – 107.6, 95% CI) for pseudotyped viruses carrying the spike protein from the D614G mutant or the Alpha, Gamma and Lambda variants, respectively (Appendix Table 3). We also observed that the geometric mean titer of the ID_50_ titers decreased by a factor of 3.05 (2.57 – 3.61, 95% CI) for the pseudotyped virus carrying the Lambda spike, 2.33 (1.95 – 2.80, 95% CI) for the Gamma spike, 2.03 (1.71 – 2.41, 95% CI) for the Alpha spike and 1.37 (1.20 – 1.55, 95% CI) for the D614G spike when compared to the Wild type spike (Figure 2A and 2B, Appendix Table 3). No correlation between sex, age, body-mass index (BMI) or smoke status and neutralizing antibody titers were observed in our study cohort (Appendix Figure 2).

**Figure 2.**
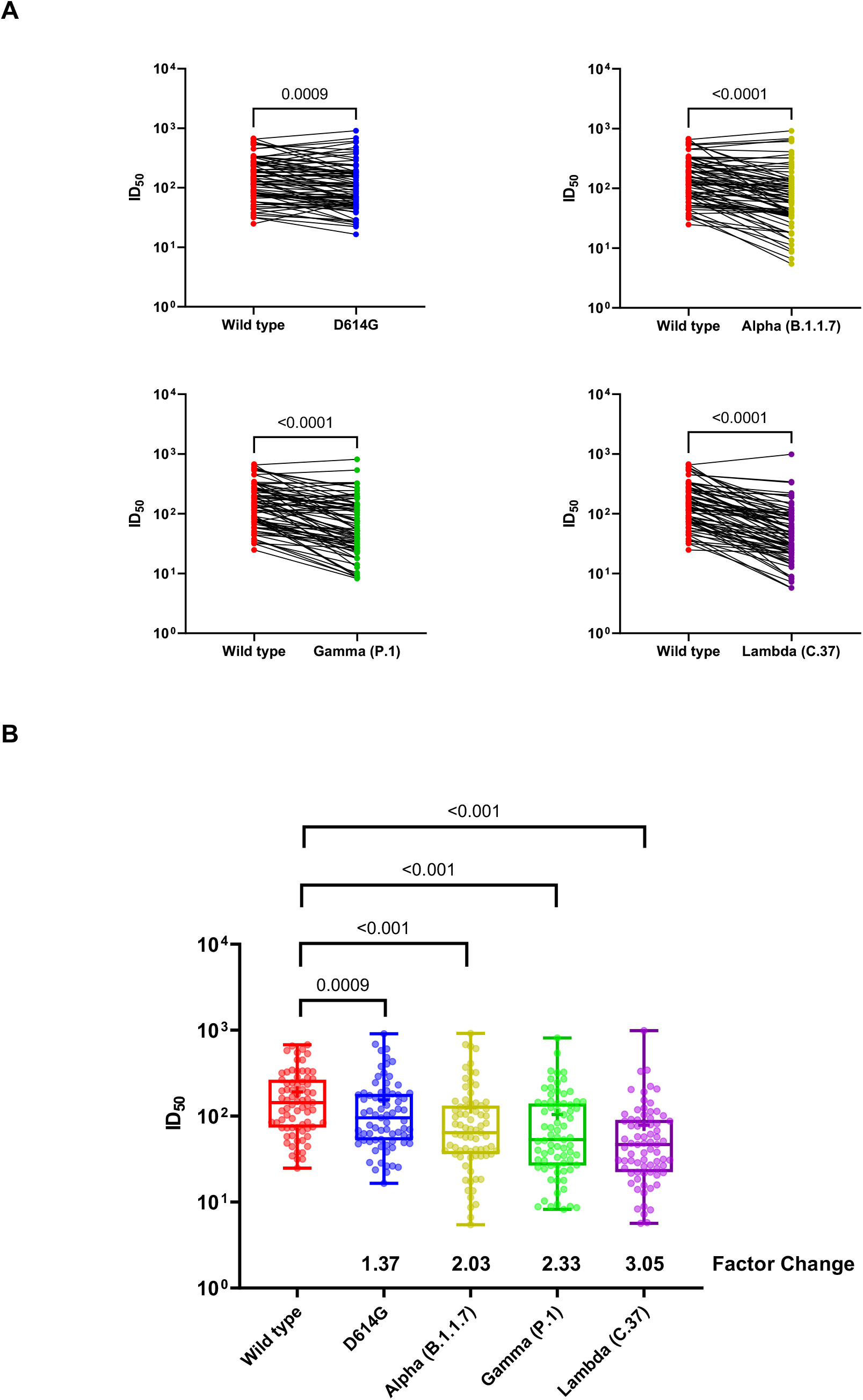
Neutralization assay using plasma samples from CoronaVac vaccinees. (A) Changes in the reciprocal 50% neutralization titer (ID_50_) in plasma samples from 75 recipients of the CoronaVac vaccine against the D614G (lineage B), Alpha (lineage B.1.1.7), Gamma (lineage P.1) and Lambda (lineage C.37) variants as compared with wild type virus. Results are shown as the difference in neutralization titers of matched samples. P values for the comparison of the ID_50_ are calculated with Wilcoxon signed-rank test. (B) Box plots indicated the median and interquartile range (IQR) of ID_50_ for each pseudotyped virus. Factor changes are shown as the difference of geometric mean titer in the ID_50_ as compared with those for the Wild type pseudotyped virus. Statistical analyses were performed using the Wilcoxon matched-pairs signed rank test.

Together, our data revealed that the spike protein of the newly recognized variant of interest Lambda, highly circulating in Chile and South American countries, carries mutations conferring increased infectivity and the ability to escape from neutralizing antibodies elicited by CoronaVac.

## DISCUSSION

High SARS-CoV-2 transmission is occurring in Chile despite an intensive vaccination campaign, which mostly relies in the inactivated virus vaccine from Sinovac Biotech and to a lesser extent in the mRNA vaccine from Pfizer/BioNTech and the non-replicative viral vector vaccines from Oxford/AstraZeneca and Cansino Biologicals. The last surge reported in the country has been dominated by the SARS-CoV-2 variants Gamma and Lambda, the former classified as a variant of concern several months ago and the latter being recently recognized as a variant of interest by the WHO. While the Gamma variant possess 11 mutations in the spike protein including those in the receptor binding domain (RBD) associated with increased ACE2 binding and infectivity (N501Y) or immune escape (K417T and E484K)^1^, the spike protein of the Lambda variant has a unique pattern of 7 mutations (Δ246-252, G75V, T76I, L452Q, F490S, D614G, T859N) from which L452Q is similar to the L452R mutation reported in the Delta and Epsilon variants^6^. The L452R mutation has been shown to confer immune escape to monoclonal antibodies (mAbs) as well as convalescent plasma^13–16^. Moreover, the L452R mutation has also been shown to increase viral infectivity^16^ and our data suggest that the L452Q mutation present in the Lambda variant might confer similar properties to those described for L452R. Interestingly, the 246-252 deletion in the N-terminal domain (NTD) of the Lambda Spike is located in an antigenic supersite^17–20^ and therefore, this deletion might also contribute to immune escape. Moreover, the F490S mutation has also been associated with an escape to convalescent sera^13^. Consistent with these antecedents, our results indicate that the spike protein of the Lambda variant confers immune escape to neutralizing antibodies elicited by the CoronaVac vaccine. Whether the Lambda variant also escapes to the cellular response shown to be elicited by CoronaVac^9^ is still unknown. We also observed that the spike protein of the Lambda variant presented increased infectivity when compared with the spike protein of the Alpha and Gamma variants, both of them with reported increased infectivity and transmissibility^21,22^. Together, our data show for the first time that mutations present in the spike protein of the Lambda variant confer escape to neutralizing antibodies and increased infectivity. The evidence presented here reinforces the idea that massive vaccination campaigns in countries with high SARS-CoV-2 circulation rates must be accompanied by strict genomic surveillance aimed to rapidly identify new viral isolates carrying spike mutations as well as studies aimed to analyze the impact of these mutations in immune escape and vaccines breakthrough.

## Data Availability

Data will be available upon request

## ACKNOWLEDGMENTS

Authors wish to thank all volunteers for participating in this study. Authors also thank to Mónica Peña (Universidad de Chile), Joseline Catrileo (Universidad de Chile), María Antonieta Núñez and the blood bank staff members (Clínica Santa María) and Margarita Gilabert and the Clinical Data Management Unit (Clínica Santa María) for technical support and sample collection. We thank Dr. Lucía Nuñez for her critical revision. ANID Chile supports the authors through Fondecyt grants N° 1190156 (R.S-R.), 1211547 (F.V.-E.) and 1181656 (A.G.).

## AUTHORS CONTRIBUTIONS

MLA, CPC, AG, FVE and RSR designed the study. MLA, LAP and AB designed mutants and performed the experiments. AG and CPC provided clinical samples. MLA, FP and FVE performed statistical analysis. CPC, FVE and RSR wrote the manuscript. FVE, AG and RSR acquired funding. All authors approved the final version of the manuscript.

## APPENDIX

**Appendix Figure 1:**
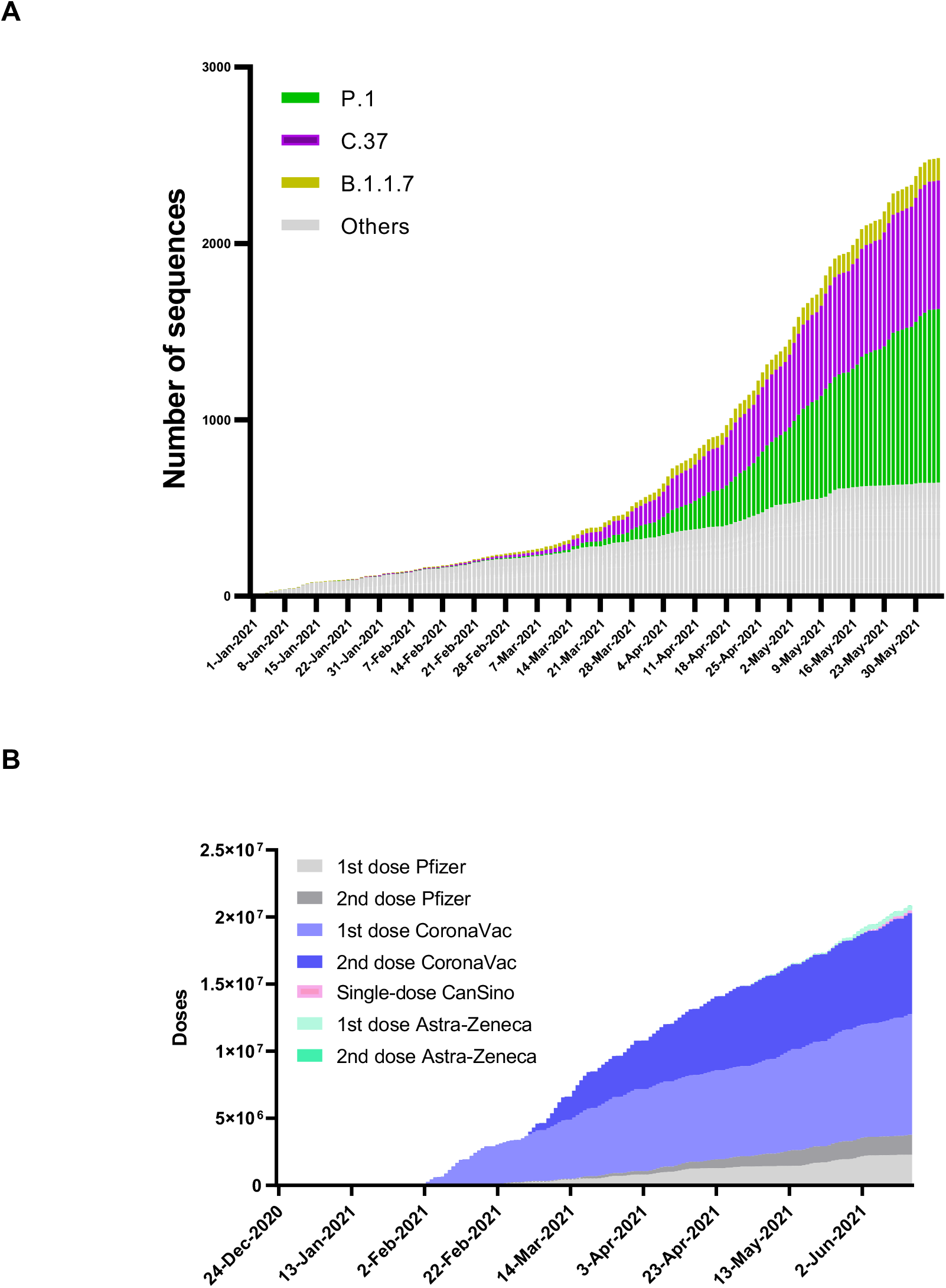
SARS-CoV-2 circulation and vaccination campaign during the first semester 2021 in Chile. (A) SARS-CoV-2 lineages circulation in Chile during the first semester 2021 according to public data deposited at https://auspice.cov2.cl/ncov/chile-global. Lineages C.37 (purple bars), P.1 (green bars) and B.1.1.7 (yellow bars) are indicated. Grey bars represent the remaining lineages. (B) Vaccine coverage during the first semester 2021 according to public data from the Ministry of Science, Technology, Knowledge and Innovation available at https://github.com/MinCiencia/Datos-COVID19 (Product 83).

**Appendix Figure 2:**
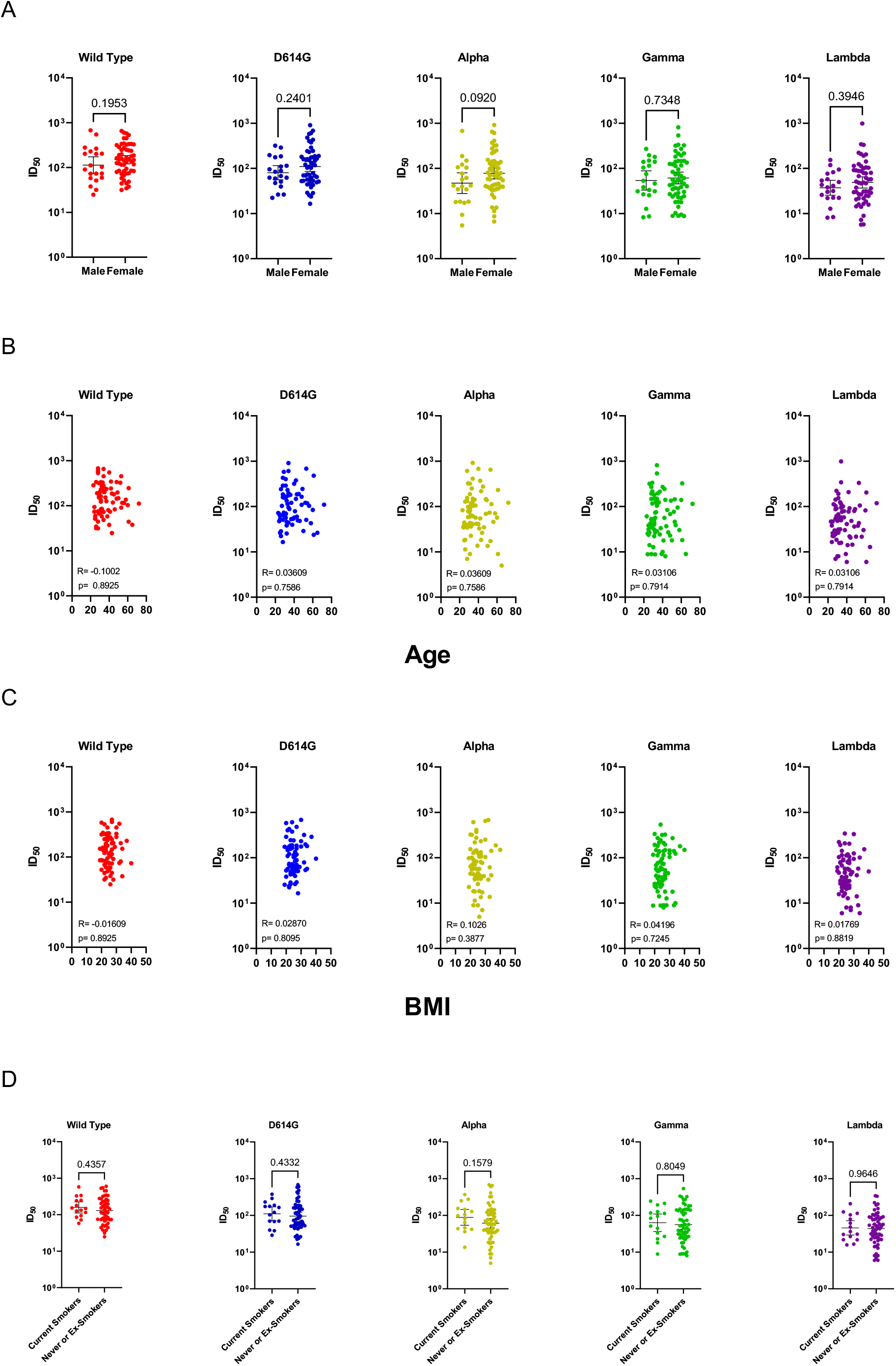
Correlation of neutralizing antibody titers against SARS-CoV-2 variants and participants’ sex, age, BMI or smoke status. Stratification neutralizing antibody titres (NAbTs) against SARS-CoV-2 variants by sex (A), Age (B), BMI (C) and smoke status (D). Neither factor significantly affects neutralizing antibody titers.

**Appendix Table 1.**
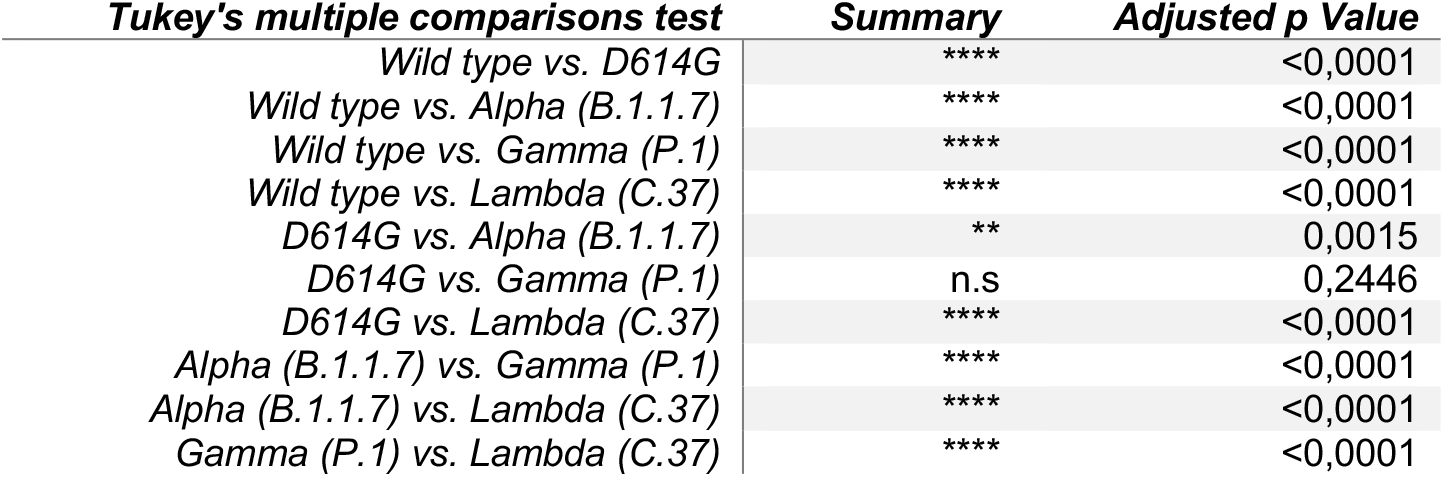
Statistical Analysis for Figure 1 at 20 ng of p24.

**Appendix Table 2.**
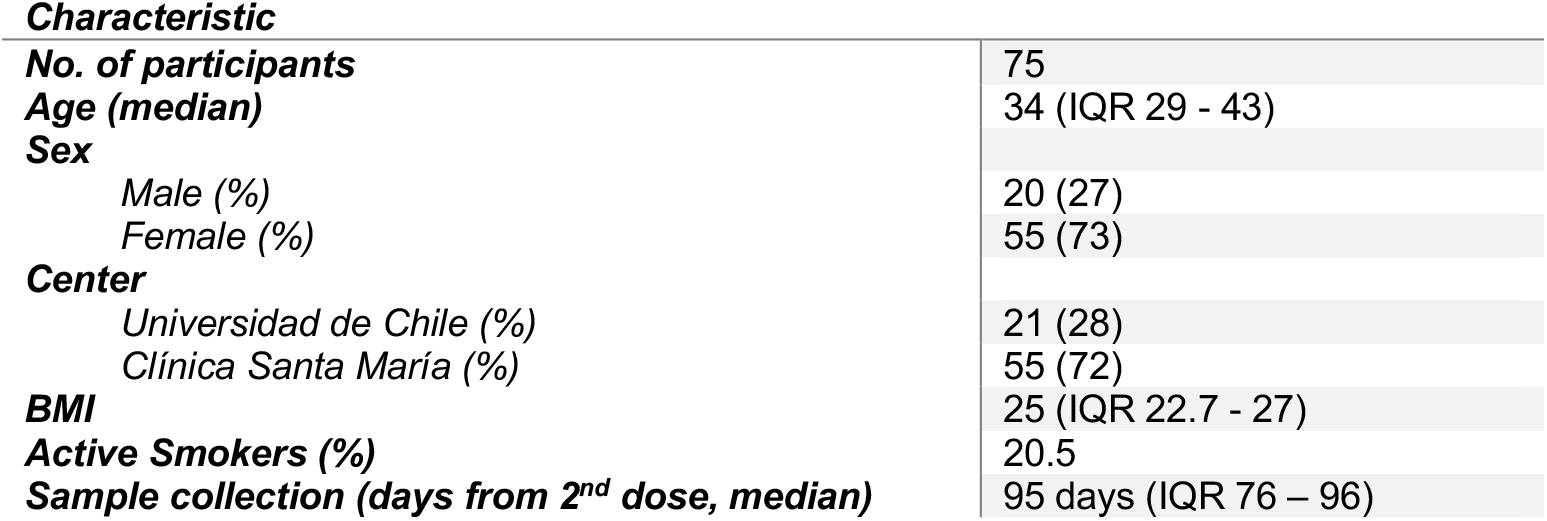
Characteristics of study subjects.

**Appendix Table 3.** Descriptive Statistics

|  | **WILD**<br>**TYPE_ID50** | **D614G_ID50** | **ALPHA_ID50** | **GAMMA_ID50** | **LAMBDA_ID50** |
| --- | --- | --- | --- | --- | --- |
| **Mean** | 191,5 | 153,9 | 124,7 | 104,6 | 78,76 |
| **std. deviation** | 158,6 | 166,2 | 167,5 | 128,4 | 125,7 |
| **std. error of mean** | 18,32 | 19,19 | 19,34 | 14,83 | 14,52 |
| **Lower 95% ci of mean** | 155 | 115,7 | 86,21 | 75,03 | 49,83 |
| **Upper 95% ci of mean** | 228 | 192,2 | 163,3 | 134,1 | 107,7 |
| **P25** | 73.6 | 52.3 | 36.1 | 26.6 | 22.3 |
| **P75** | 265.1 | 181.4 | 132.6 | 140.5 | 90.32 |
| **IQR** | 191.48 | 129.03 | 96.45 | 113.84 | 67.96 |
| **P50** | 143.9 | 95.68 | 64.11 | 53.22 | 46.66 |

**Descriptive Statistics of differences in the neutralization titers relative to the wild type**
|  | **wild**<br>**type_id50/d614g**<br>**_id** | **wild**<br>**type_id50/alpha**<br>**_id** | **wild**<br>**type_id50/gamma**<br>**_id** | **wild**<br>**type_id50/lambda**<br>**_id** |
| --- | --- | --- | --- | --- |
| **Mean** | 37,5 | 66,7 | 86,9 | 112,7 |
| **std. deviation** | 14,3 | 15,1 | 14,8 | 15,8 |
| **std. error of mean** | 124,0 | 13,7 | 128,5 | 136,8 |
| **Lower 95% ci of mean** | 9,0 | 36,6 | 57,3 | 81,2 |
| **Upper 95% ci of mean** | 66,06 | 96,8 | 116,44 | 114,1 |
| **Geometric mean** | 1,37 | 2,035 | 2,34 | 3,05 |
| **Geometric sd factor** | 1,717 | 2,108 | 2,193 | 2,08 |

## REFERENCES

1. Harvey WT, Carabelli AM, Jackson B, et al. SARS-CoV-2 variants, spike mutations and immune escape. Nat Rev Microbiol 2021;19(7).

2. Gupta RK. Will SARS-CoV-2 variants of concern affect the promise of vaccines?. Nat. Rev. Immunol. 2021;21(6):340–1.

3. Krause PR, Fleming TR, Longini IM, et al. SARS-CoV-2 Variants and Vaccines. N Engl J Med 2021

4. COVID-19 Weekly Epidemiological Update Global overview. Availble at: https://www.who.int/publications/m/item/weekly-epidemiological-update-on-covid-19---15-june-2021

5. Nextstrain / community / quipupe / C37_lineage. Available at: https://nextstrain.org/community/quipupe/C37_lineage

6. Novel sublineage within B.1.1.1 currently expanding in Peru and Chile, with a convergent deletion in the ORF1a gene (Δ3675-3677) and a novel deletion in the Spike gene (Δ246-252, G75V, T76I, L452Q, F490S, T859N) - SARS-CoV-2 coronavirus / nCoV-2019 Genomic Epidemiology - Virological Available at: https://virological.org/t/novel-sublineage-within-b-1-1-1-currently-expanding-in-peru-and-chile-with-a-convergent-deletion-in-the-orf1a-gene-3675-3677-and-a-novel-deletion-in-the-spike-gene-246-252-g75v-t76i-l452q-f490s-t859n/685

7. Chilean Ministry of Health Vaccine Report. Available at: https://informesdeis.minsal.cl/SASVisualAnalytics/?reportUri=%2Freports%2Freports%2F9037e283-1278-422c-84c4-16e42a7026c8&sectionIndex=0&sso_guest=true&reportViewOnly=true&reportContextBar=false&sas-welcome=false

8. Muena NA, García-Salum T, Pardo-Roa C, et al. Long-lasting neutralizing antibody responses in SARS-CoV-2 seropositive individuals are robustly boosted by immunization with the CoronaVac and BNT162b2 vaccines. medRxiv Prepr Serv Heal Sci 2021

9. Bueno SM, Abarca K, González PA, et al. INTERIM REPORT: SAFETY AND IMMUNOGENICITY OF AN INACTIVATED VACCINE AGAINST SARS-COV-2 IN HEALTHY CHILEAN ADULTS IN A PHASE 3 CLINICAL TRIAL 2 3 Brief Title: CoronaVac03CL Phase 3 Interim Analysis of Safety and Immunogenicity. medRxiv 2021;2021.03.31.21254494.

10. Wang G-L, Wang Z-Y, Duan L-J, et al. Susceptibility of Circulating SARS-CoV-2 Variants to Neutralization. N Engl J Med 2021;384(24):2354–6.

11. Khoury DS, Cromer D, Reynaldi A, et al. Neutralizing antibody levels are highly predictive of immune protection from symptomatic SARS-CoV-2 infection. Nat Med 2021;1–7.

12. Beltrán-Pavez C, Riquelme-Barrios S, Oyarzún-Arrau A, et al. Insights into neutralizing antibody responses in individuals exposed to SARS-CoV-2 in Chile. Sci Adv 2021;7(7).

13. Liu Z, VanBlargan LA, Bloyet LM, et al. Identification of SARS-CoV-2 spike mutations that attenuate monoclonal and serum antibody neutralization. Cell Host Microbe 2021;29(3):477-488.e4.

14. Li Q, Wu J, Nie J, et al. The Impact of Mutations in SARS-CoV-2 Spike on Viral Infectivity and Antigenicity. Cell 2020;182(5):1284-1294.e9.

15. Greaney AJ, Starr TN, Gilchuk P, et al. Complete Mapping of Mutations to the SARS-CoV-2 Spike Receptor-Binding Domain that Escape Antibody Recognition. Cell Host Microbe 2021;29(1):44-57.e9.

16. Deng X, Garcia-Knight MA, Khalid MM, et al. Transmission, infectivity, and neutralization of a spike L452R SARS-CoV-2 variant. Cell 2021;184(13):3426–3437.e8.

17. Venkatakrishnan AJ, 2+ A, Lenehan P, et al. Antigenic minimalism of SARS-CoV-2 is linked to surges in COVID-19 community transmission and vaccine breakthrough infections. medRxiv 2021;2021.05.23.21257668.

18. McCallum M, De Marco A, Lempp FA, et al. N-terminal domain antigenic mapping reveals a site of vulnerability for SARS-CoV-2. Cell 2021;184(9):2332-2347.e16.

19. Cerutti G, Guo Y, Zhou T, et al. Potent SARS-CoV-2 neutralizing antibodies directed against spike N-terminal domain target a single supersite. Cell Host Microbe 2021;29(5):819-833.e7.

20. McCarthy KR, Rennick LJ, Nambulli S, et al. Recurrent deletions in the SARS-CoV-2 spike glycoprotein drive antibody escape. Science 2021;371(6534):1139–42.

21. Faria NR, Mellan TA, Whittaker C, et al. Genomics and epidemiology of the P.1 SARS-CoV-2 lineage in Manaus, Brazil. Science 2021;372(6544):815–21.

22. Davies NG, Abbott S, Barnard RC, et al. Estimated transmissibility and impact of SARS-CoV-2 lineage B.1.1.7 in England. Science 2021;372(6538):eabg3055.

